# Modeling rod and cone photoreceptor cell survival *in vivo* using optical coherence tomography

**DOI:** 10.1101/2022.11.21.22281626

**Authors:** S. Scott Whitmore, Adam P. DeLuca, Jeaneen L. Andorf, Justine L. Cheng, Mahsaw Mansoor, Christopher R. Fortenbach, D. Brice Critser, Jonathan F. Russell, Edwin M. Stone, Ian C. Han

## Abstract

Many retinal diseases involve the loss of light-sensing photoreceptor cells (rods and cones) over time. The severity and distribution of photoreceptor loss varies widely across diseases and affected individuals, so characterizing the degree and pattern of photoreceptor loss can clarify pathophysiology and prognosis. Currently, *in vivo* visualization of individual photoreceptors requires technology such as adaptive optics, which has numerous limitations and is not widely used. By contrast, optical coherence tomography (OCT) is nearly ubiquitous in daily clinical practice given its ease of image acquisition and detailed visualization of retinal structure. However, OCT cannot resolve individual photoreceptors, and no OCT-based method exists to distinguish between the loss of rods versus cones. Here, we present a computational model that quantitatively estimates rod versus cone photoreceptor loss from OCT. Using histologic data of human photoreceptor topography, we constructed an OCT-based reference model to simulate outer nuclear layer thinning caused by differential loss of rods and cones. The model was able to estimate rod and cone loss using *in vivo* OCT data from patients with Stargardt disease and healthy controls. Our model provides a powerful new tool to quantify photoreceptor loss using OCT data alone, with potentially broad applications for research and clinical care.

## Introduction

Photoreceptor loss is a shared feature of many retinal diseases that cause vision loss, including common conditions such as age-related macular degeneration and rare conditions such as Stargardt disease. The two main subtypes of human photoreceptor cells are rods and cones. Differential death of rods versus cones can produce distinct visual symptoms even among individuals with the same disease. These differences arise, in part, from the contrasting physiology and function of the two cell types, but also from their differential distribution in the retina. Cone photoreceptors are tightly packed in the fovea and progressively less concentrated more anteriorly, whereas rods predominate in the peripheral retina, reach their greatest density in a ring at the eccentricity of the optic nerve, and are absent from the very center of the fovea ^1^.

While some psychometric tests can differentiate rod versus cone loss (for example, Ref.^2^), current imaging methods for visualizing and quantifying rod and cone loss have significant limitations. For example, adaptive optics (AO) techniques enable visualization of individual cones^3-8^ and sometimes rods^5^. However, commercial AO systems are not widely available, image only a narrow area of the retina at a time, and are technically challenging to use.

By contrast, commercially available OCT systems have been widely used in routine clinical practice for almost two decades, providing a wealth of longitudinal data for both qualitative and quantitative assessment of changes over time. The outer nuclear layer on OCT scans represents photoreceptor nuclei. Nevertheless, OCT can only capture bulk trends of total photoreceptor survival since these systems lack the requisite resolution to differentiate between single cells. No methods currently exist to estimate the proportions of surviving rods versus cones from OCT images. In lieu of individual cell counts, even approximations of rod and cone survival by OCT would be useful for assessing disease progression in clinical trials or routine care.

In this study, we developed a strategy for extracting and assessing the relative survival of rods and cones from *in vivo* OCT datasets. Our model leverages previously published histologic data on human photoreceptor topography^1^ to relate the thickness of the outer nuclear layer on OCT to the expected survival of rods and cones. To demonstrate applicability of the model to real-world clinical data, we applied this model to OCTs from a cohort of patients with autosomal recessive Stargardt disease (STGD1)^9^ to estimate the proportional survival of rods and cones.

## Methods

Our method compares retinal layer thickness data derived from patient OCTs to retinal layer thickness data generated from a computational reference model. The computational model simulates a spectrum of rod and cone survival scenarios, relating the proportion of surviving rods versus cones to changes in the combined thickness of the Henle fiber layer (HFL), outer nuclear layer (ONL), and myoid zone (MZ). For simplicity throughout the paper, we refer to the combined HFL-ONL-MZ layers as the ONL.

### 1. Building the reference model

#### 1.1 Imaging for the reference model

As proof of concept, we imaged the maculas of an unaffected, young adult control patient using a Heidelberg Spectralis OCT (Heidelberg Engineering, Heidelberg, Germany). Each volume contained 61 B-scans. We exported these volumes from Heidelberg Explorer as VOL files. For each volume, we segmented 11 retinal layers using the Iowa Reference Algorithms/OCTExplorer (ver. 3.8.0; Retinal Image Analysis Lab, Iowa Institute for Biomedical Imaging, Iowa City, IA)^10,11^. We adopted the nomenclature proposed in Ref.^12^ to identify the layers segmented by the Iowa Reference Algorithms. We located the fovea and the optic nerve within these volumes to serve as common landmarks for registering cell density data to scans.

#### 1.2 Data Integration

We used *heyexr* (ver 0.0.0.9000; https://github.com/barefootbiology/heyexr), our custom software package, to import the OCT VOL files and segmentation files into R (ver 4.1.3^13^). We registered these volumes to published densities of photoreceptor cells from Curcio and coworkers’ landmark 1990 paper on photoreceptor densities (Ref.^1^; original data available at https://christineacurcio.com/PRtopo/). We interpolated the cell densities at every A-scan using the akima function from the *akima* package (ver. 0.6-3.3^14^) for R. The *akima* method creates a continuous, smooth interpolation between irregularly spaced sampling points, such as those used by Curcio and coworkers, while preserving values at the sampling points. Although the absolute number of photoreceptor cells can vary widely across individuals, the relative proportion of rods and cones for a given topographic location is very similar. As such, we converted the cell density values to proportions of cells by dividing the density of rods by the total density of both cell types. The completed reference model for each eye comprises: (a) the original OCT volume scan; (b) the layer segmentation surfaces; and (c) the expected proportion of rods and cones at every A-scan in the volume.

### 2. Deriving proportional thickness values from the reference model

#### 2.1 Assumptions

We assumed that the thickness of the outer retina is proportional to the topographic composition of surviving rod and cone photoreceptor cells. As rods and/or cones die, the thickness of the outer retina will decrease relative to the contribution each cell type makes to total retinal thickness at that location. In eyes unaffected by disease, we assumed that 100% of photoreceptors survive. We refer to the retinal layer thickness of eyes unaffected by disease as the *normal thickness*. Thus, the normal thickness of the ONL at any given location implies survival of all photoreceptor cells at that location. We assumed that the distribution and density of photoreceptor cells in a normal eye equals the average distribution and density of photoreceptor cells reported in Ref.^1^, that is, the same data included in the reference model. We also assumed that individual rod and cone cells contribute equally to ONL thickness for any given topographic location.

#### 2.2 Mathematical framework

Given these assumptions, the observed thickness (*T*_*observed*_) of the ONL will be proportional to the normal thickness (*T*_*normal*_) times the proportion of surviving rods and cones (*P*_*thickness*_):

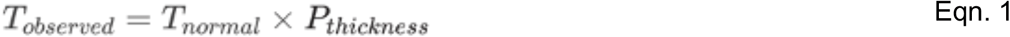

The proportional thickness (*P*_*thickness*_) is a function of the relative composition of cones (*P*_*cones*_) and rods (*P*_*rods*_) at a location and the proportion of surviving cones (*S*_*cones*_) and rods (*S*_*rods*_) at that location:

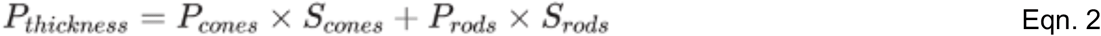

Since rods and cones are the only cell types in the outer retina (apart from the negligible extension of the Müller glia cells), the relative proportion of rods plus the relative proportion of cones equals 1 (100%). Rearranging, we get:

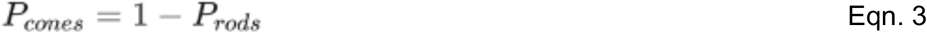

We substitute this term into **Equation 2** and get:

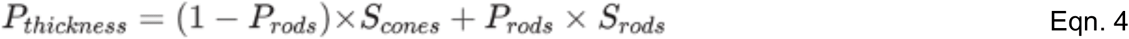

The surviving cones and surviving rods become parameters in the model. By changing the proportion of these two parameters (denoted by asterisks, *), we can compute a simulated proportional thickness:

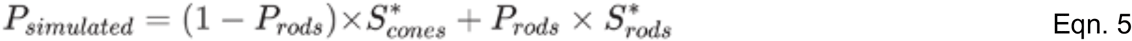

and a new simulated thickness:

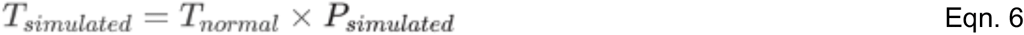

With this mathematical framework and the reference model, **Equations 5** and **6** let us simulate a set of thickness values for the ONL. By varying the *S*_*cones*_ and *S*_*rods*_ terms from 0 to 1, we generated a range of hypothetical thickness values for the ONL. From this set of simulated thickness data, we constructed corresponding segmentation surfaces and *en face* layer thickness maps.

**Equation 4**, solved for a single region of the retina, will not produce a unique solution to the proportion of rods and cones, since there are two output variables but only one equation. If a second region is added, which has a different expected proportion of rods and cones, then the system of equations will have a unique solution for the global estimates of surviving rods and cones.

#### 2.3 Layer thickness within the center subfield and outer ring of the ETDRS grid

From our simulated thickness maps, we computed the average layer thickness in the center subfield and outer ring of the ETDRS grid. The ETDRS grid is a familiar clinical tool for standardized assessment of pathological features in the macula. The grid is composed of nine subfields: a subfield centered on the fovea, four inner subfields arranged in a ring, and four outer subfields arranged in a ring^15^. Conveniently, the annular arrangement of these rings reflects the photoreceptor organization of the macula: the center subfield is predominantly cones; the inner ring includes the transition from cone-dominated retina to rod-dominated retina; and the outer ring is predominantly rods. While the peak cone density can vary markedly by individuals^1,7^, the center subfield is reliably a cone-predominant zone with a consistent proportion of cones versus rods across individuals. Similarly, the exact location of the transition zone within the inner ring from cone-dominated retina to rod-dominated retina varies widely, but the relative proportions of rods and cones is more consistent between individuals in the outer ring. For these reasons, we analyzed only the center subfield and outer ring. The outer ring is typically divided into four subfields each; however, we chose to treat the ring as a unit. This choice made the analysis invariant to the rotation of the retina: we did not need to register the positions of the fovea and optic nerve to a common orientation when computing average thickness.

### 3. Comparing observed clinical data to the simulated reference data

To demonstrate the utility of this model for analysis of clinical data, we used estimates of ONL thickness from our previous publication on autosomal recessive Stargardt disease (STGD1), which included OCT volume scans from 50 STGD1 patients and 40 unaffected controls^9^. The research was approved by the Institutional Review Board at the University of Iowa, adhered to the tenets of the Declaration of Helsinki, and was conducted in accordance with regulations set forth by the Health Insurance Portability and Accountability Act. Informed consent was obtained for study participation. For participants under the age of 18 years, informed consent was obtained from a parent and/or legal guardian. We chose Stargardt disease because photoreceptor loss occurs within the macula and can affect cones and rods to varying degrees across individuals. Briefly, volume OCTs were collected under various imaging protocols (20 degrees × 20 degrees to 30 × 20 degrees, with 512, 768 or 1024 (horizontal) × 496 (vertical) pixel density, and comprised of 19, 25, 31, 37, 47, or 49 B-scans). Follow-up scans were registered to baseline scans using TruTrack Active Eye Tracking. Follow-up scans were taken at least 12 months after baseline scanning. Layer segmentation was performed using the Iowa Reference Algorithms. Segmentation surfaces were corrected by two graders (C.R.F, J.L.C). Graders removed scans which could not be used for analysis. The average segmented layer thickness was computed within the center subfield and outer ring of the ETDRS grid. The average thickness of each subfield was modeled as a function of years since baseline and STGD1 or control status, using linear mixed effects models in R (nlme ver. 3.1^16,17^). Specifics on statistical modeling, demographics of the patient and control groups, and exclusion criteria are detailed in Ref.^9^.

## Results

As proof of concept, we built a reference model consisting of (a) the OCT scan of an eye unaffected by disease; (b) expected cell densities of rod and cone photoreceptors; and (c) segmentation surfaces for the retinal sublayers, as shown in **Figures 1 and 2. Figure 1A** shows the *en face* projection of OCT data that we used to build the reference model. The subject was a male in their 30s with no known retinal disease. Using the fovea and optic nerve of each eye as landmarks, we registered cell density estimates from Ref.^1^. In that study, the densities of rods and cones were sampled in a spiral pattern (explained in Ref.^18^) in seven donor eyes, averaged, registered to the fovea and optic nerve of an idealized anatomic model eye^19^, and reported for a left eye (OS). Since the dimensions of OCT scans are recorded in millimeters, we converted the original spherical coordinates (longitude and colatitude centered on the fovea) of the cell density data^1^ to the planar coordinates (millimeters from the fovea along the vertical and horizontal meridians) of the OCT. The anatomic model adopted by Curcio and coworkers assumes that the fovea and the optic disc lie along the same horizontal meridian (0° degrees longitude)^1^; however, the fovea typically lies slightly below the horizontal meridian passing through the optic disc. Furthermore, the relative position of these two anatomic landmarks may vary based on the positioning of the subject during OCT imaging, as illustrated in **Figure 1A**. To account for differences in the position of the fovea and optic nerve within an OCT scan, we registered the cell density sampling points to the optic nerve and fovea in the control OCTs and flipped the points along the vertical axis for the right eye (OD) (**Figure 1B**). The published sampling points of photoreceptors, however, do not align with the A-scan positions of the OCT (that is, every pixel position in the *en face* projection shown in **Figure 1A**). To account for this difference, we generated cell density maps by smoothly interpolating the average cell densities of cones (**Figure 1C**) and rods (**Figure 1D**) across every A-scan position in the OCTs.

**Figure 1:**
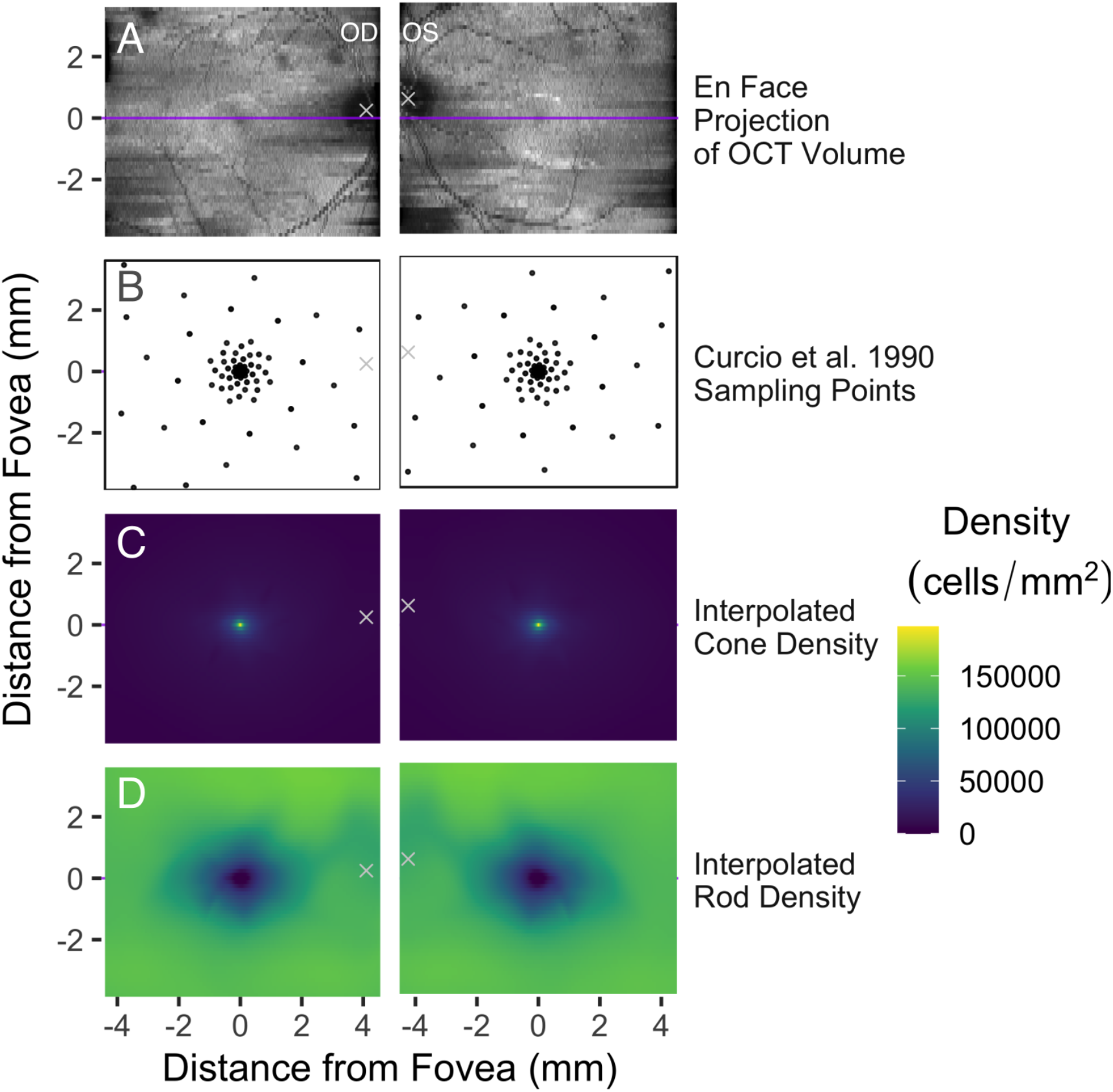
Registering OCT volumes to publicly available photoreceptor cell density data. (**A**) OCT scans of right (OD) and left (OS) eyes from a healthy adult control, represented as an *en face* projection (that is, the sum of voxel intensities in each A-scan). The coordinate space is centered on the fovea, and the center of the optic nerve is marked by a white ‘x’. The purple horizontal line indicates the B-scan in each eye which passes through the fovea (see Figure 2). The *y*-axis of each pair of panels is aligned to the fovea, accounting for the slight offset between the left and right columns. (**B**) Sampling points from Ref.^1^ (described in Ref.^18^) which fall within the extent of the OCT. Points have been rotated and scaled to align with the fovea and optic nerve in each OCT. (**C-D**) Density of rod photoreceptor cells (**C**) and cone photoreceptor cells (**D**) interpolated for each A-scan position using Akima interpolation^14^.

**Figure 2.**
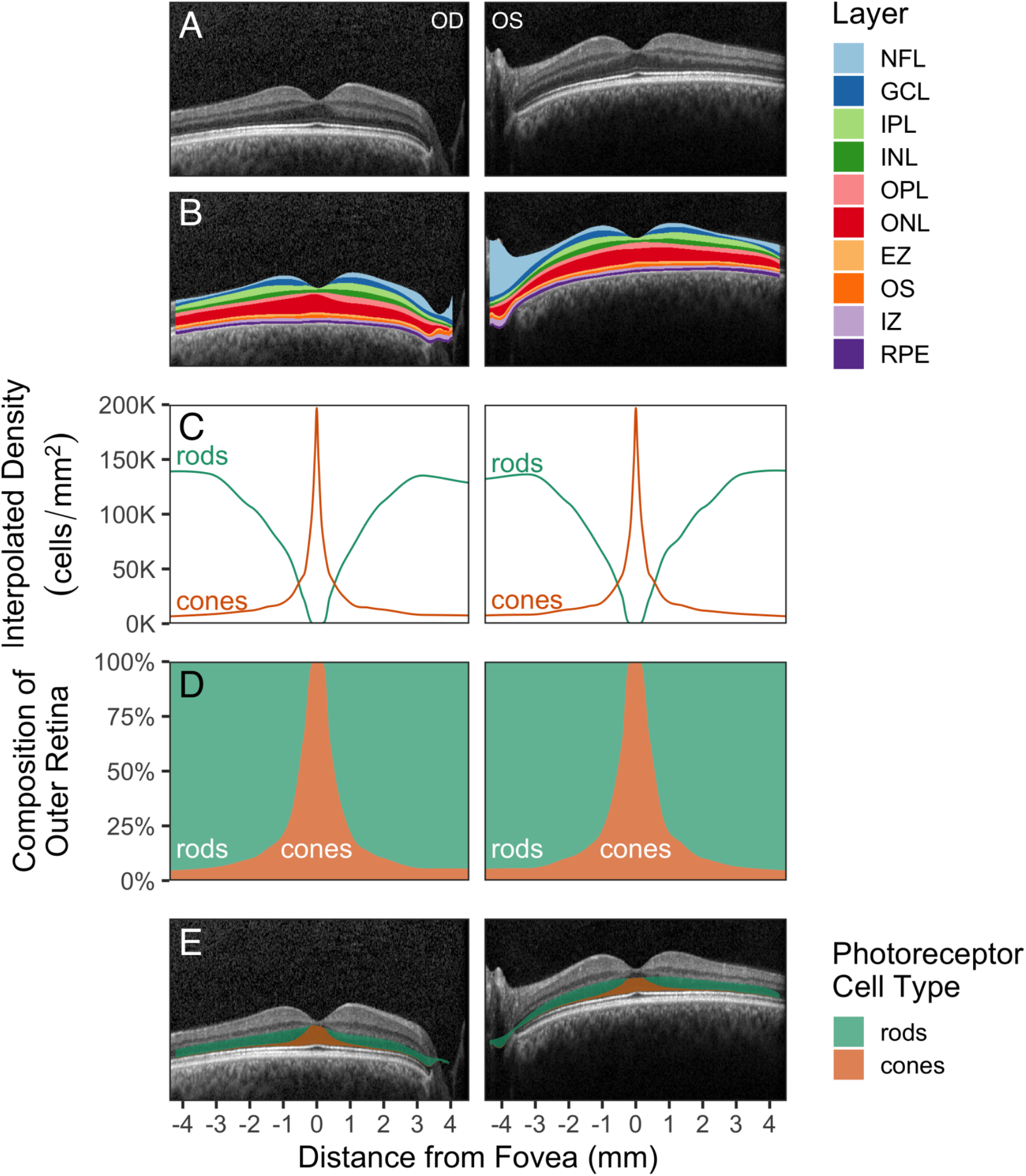
The reference model relates ONL thickness to expected proportions of rods and cones. (**A**) Central B-scans from the control eyes shown in Figure 1A. (**B**) Segmentation surfaces identified by Iowa Reference Algorithms correspond to 10 layers. The red layer comprises the Henle fiber layer (HFL), the outer nuclear layer (ONL), and the myoid zone (MZ). (**C**) Interpolated photoreceptor densities registered to the central B-scan. (**D**) Proportional composition of rods and cones across the B-scan. (**E**) The expected proportional composition of the ONL. Abbreviations: NFL, nerve fiber layer; GCL, ganglion cell layer; IPL, inner plexiform layer; INL, inner nuclear layer; OPL, outer plexiform layer; ONL, Henle fiber layer, outer nuclear layer, and myoid zone; EZ, ellipsoid zone; OS, outer segments; IZ, interdigitation zone; RPE, retinal pigment epithelium.

Next, we segmented the retinal layers of the control OCT using publicly available software^10,11^. **Figure 2A** shows the central B-scan for each eye without segmentation, and **Figure 2B** shows the central B-scan overlaid with the segmented layers.

The aim of the reference model is to relate observed retinal thickness to the expected proportions of surviving rods and cones. From the expected photoreceptor densities (**Figure 2C**), we computed the expected proportion of rods and cones at each A-scan (**Figure 2D**). Assuming that the rod cell bodies and cone cell bodies make equal contributions to the thickness of the ONL, we can visualize the thickness of the ONL as coming from two components, with each component proportional to the percentage of rods and cones at a given eccentricity from the fovea (**Figure 2E**).

The assembled reference model simulates the change in thickness that would occur if rods or cones were lost. **Figure 3A** shows thickness maps of the ONL for the right eye. The map in the upper right corner of **3A** corresponds to the segmented layer thickness in the control eye. We assumed that all photoreceptors are living in the control eye. The other thickness maps in **3A** are simulated from the reference model, showing the change in thickness as a function of the independent loss of either cell type. Cones comprise the major photoreceptor cell type of the center subfield of the ETDRS grid^15^, whereas rods comprise the major photoreceptor cell type of the outer ring (see section 2.3 of Methods). **Figure 3B** shows the center subfield and outer ring overlaid on the thickness map from the upper right corner of **Figure 3A**. For each of the thickness maps shown in **Figure 3A**, we computed the average thickness of the ONL for the center and outer subfields and plotted these subfields along their respective cell survival parameters (**Figure 3C**).

**Figure 3.**
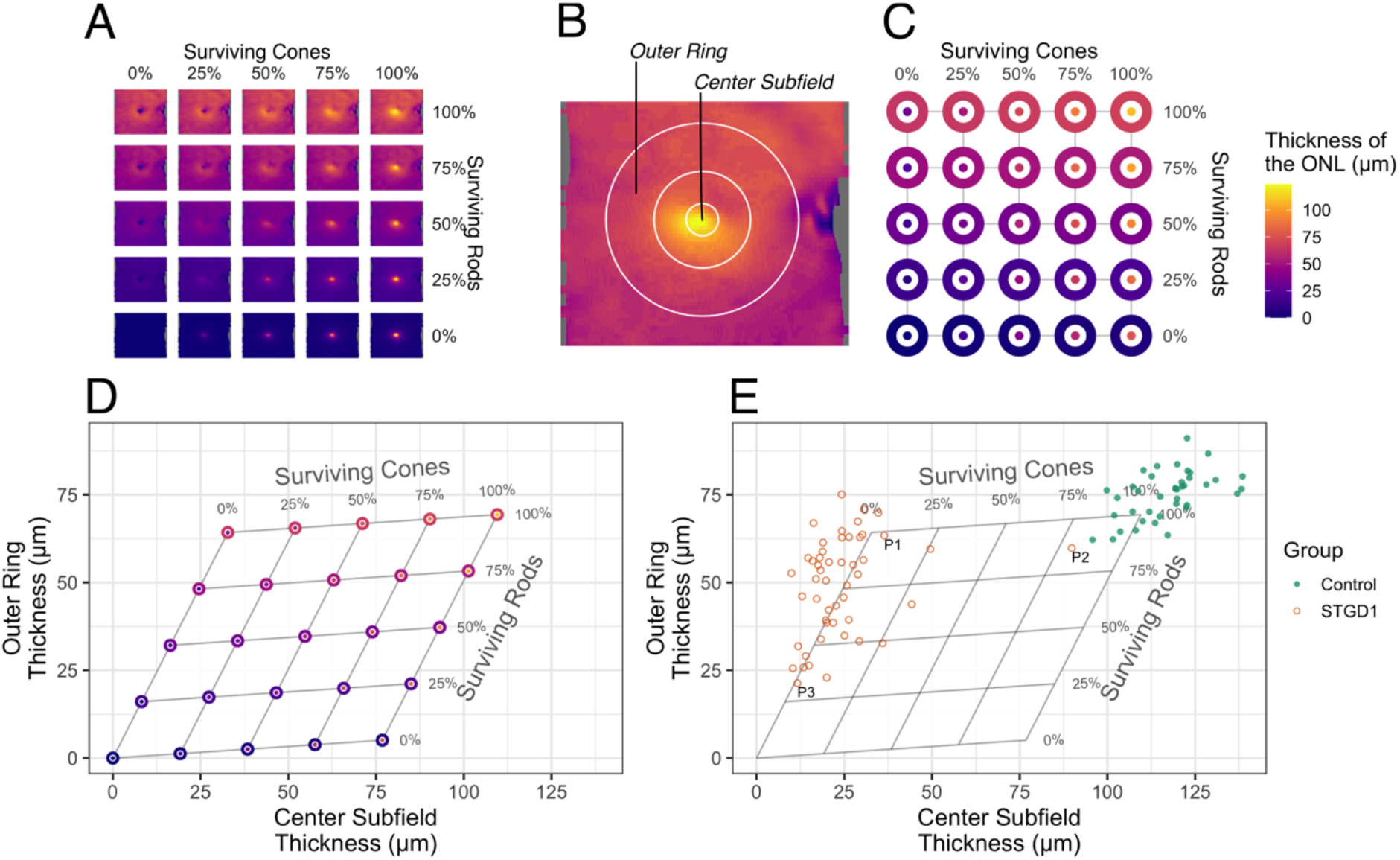
The reference model estimates rod versus cone survival based on ONL thickness. (**A**) *En face* thickness maps of the ONL from the right control eye used for the reference model. (**B**) Center subfield and outer ring of the ETDRS grid registered to the observed thickness map from the control used as input for the simulation. This panel is the same data represented in the upper right corner of Figure 3A. **(C)** Mean thickness in the center subfield and outer ring of the ETDRS grid for the observed (upper right corner) and simulated (all other locations) loss of rod and cone photoreceptor cells. (**D**) Cell survival mapped onto ETDRS subfield thickness coordinates. **(E)** ONL thickness values for 50 autosomal recessive Stargardt disease (STGD1) patients and 40 controls at baseline visit. Each point represents the fitted mean at baseline for each person (two eyes and two visits; visits at least 1 year apart; fitted using linear mixed effects models^9^). The three patients labeled (P1, P2, P3) are shown in Figure 4.

Since clinicians are most familiar with considering the thickness of the retina rather than proportions, we transformed the reference model from a graph of proportional survival (**Figure 3C**) to a graph of regional thickness (**Figure 3D**). Within **Figure 3D**, the relationship between thickness and cellular composition becomes apparent in the skewed arrangement of points. (If the center subfield was composed entirely of cones and the outer ring composed entirely of rods, then the points in **Figure 3D** would instead form a rectangle.) In other words, the skewed graph reflects that these two regions are skewed mixes of both cell types, with the center subfield predominantly composed of cones and the outer ring predominantly composed of rods. Thus, this model translates *in vivo* measurements of regional ONL thickness into estimated proportions of surviving rods and cones.

To illustrate the clinical utility of this model, we plotted ONL thickness values from 50 patients with autosomal recessive Stargardt disease (STGD1) and 40 controls (**Figure 3E**), the subset of the cohort we published previously^9^ which have thickness estimates for the center subfield and outer ring of the ETDRS grid. STGD1, as a genetic condition, affects both eyes similarly. For this reason, in our previous paper we used mixed effects modeling to estimate a per-person thickness and rates of disease progression^9^. Thus, each point in **Figure 3E** represents the per-person thickness at the baseline visit, estimated using linear mixed effects models based on one or two eyes and two visits per eye^9^. Separate models were fit for the center and outer subfields.

To highlight the ability of the model to differentiate between patients based on degrees of rod and cone loss, we selected three patients (P1-P3, **Figure 3E**) with different estimates of photoreceptor loss. **Figure 4** illustrates the correspondence between their B-scan data and estimated rod versus cone proportions. As shown in the first example, the model detected not only the obvious severe central cone loss in P1 but also subtle parafoveal rod loss, which was not easy to perceive qualitatively on the B-scan (**Figure 4A**). As shown for P2, mild loss of the parafoveal rods was clinically apparent due to disruption of the outer retinal layers (e.g., ellipsoid zone band), which draws attention to pathological thinning. However, the model also detected mild cone loss, which was not as obvious on initial qualitative assessment given the preservation of the subfoveal outer retinal layers on B-scan (**Figure 4B**). In P3, severe loss of both cones and rods was readily apparent (**Figure 4C**), and the model provided a quantitative estimate of the degree of loss for each cell type.

**Figure 4.**
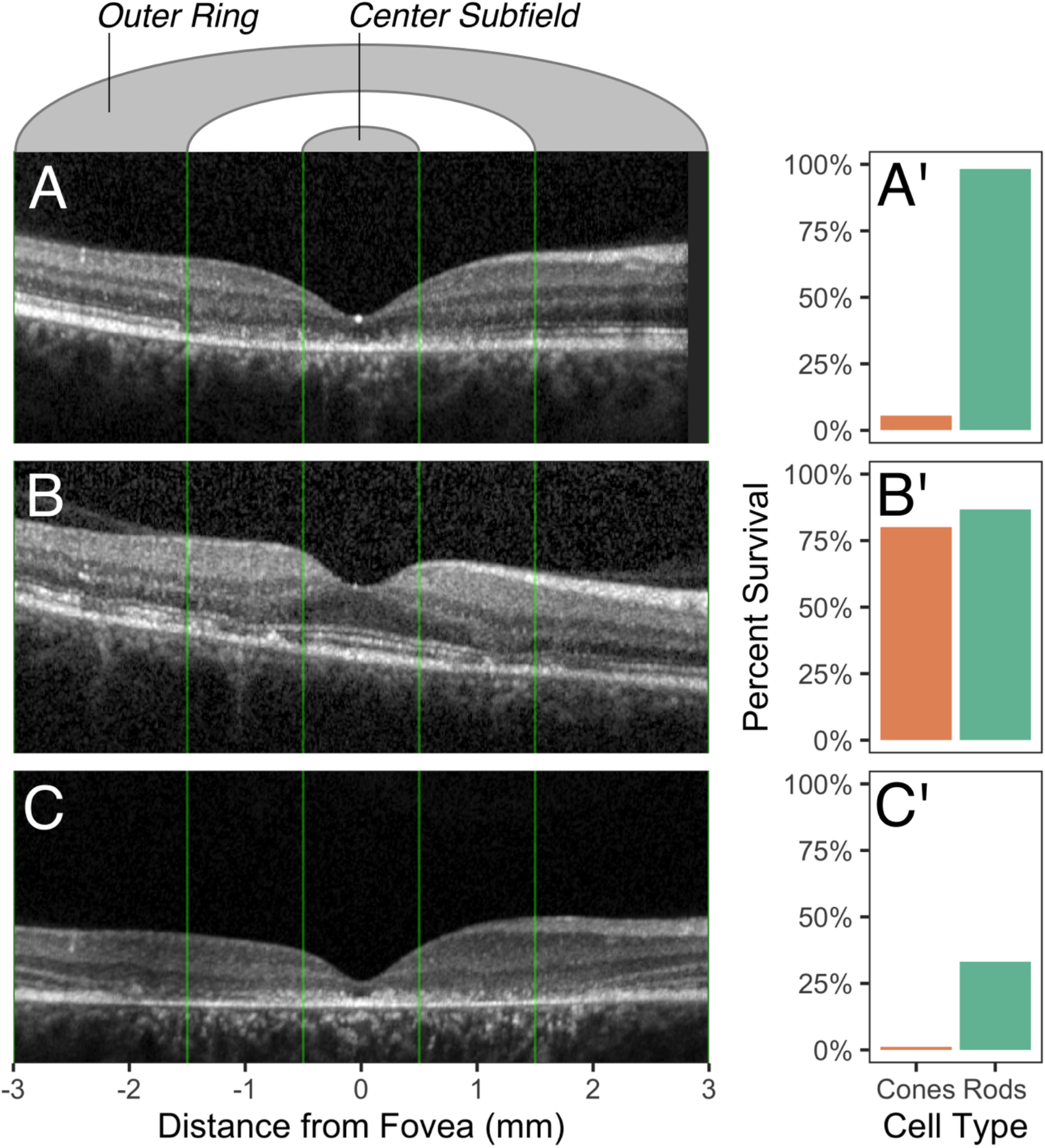
Estimated photoreceptor survival and foveal B-scans for three STGD1 patients. Patient P1 (female, in their third decade of life, best corrected visual acuity [BCVA] 20/100 OD) shows reduced foveal thickness with near normal parafoveal thickness (**A**), corresponding to severe cone loss and minimal rod loss (**A’**). Patient P2 (female, in their sixth decade of life, BCVA 20/20 OD) shows relative subfoveal preservation of the outer retinal layers with patches of parafoveal thinning (**B**), corresponding to slightly reduced cones and rods (**B’**). Patient P3 (female, in their second decade of life, BCVA 10/160 OD) shows widespread outer retinal loss (**C**), corresponding to nearly complete loss of cones, with some rods remaining (**C’**). The estimates of photoreceptor survival are computed with respect to the reference model. B-scans have been cropped to the extent of the ETDRS outer ring. Each patient is labeled in Figure 3E.

## Discussion

In this paper, we present a new computational method for utilizing *in vivo* retinal layer thickness data captured by optical coherence tomography (OCT) to estimate the degree of rod and cone photoreceptor loss. Our model is registered to photoreceptor density data from landmark literature on human photoreceptor topography^1^ and takes advantage of freely-available software^10,11^ that performs retinal sublayer segmentation to isolate the ONL thickness for quantification of photoreceptor survival. Rather than attempting to quantify the absolute density of photoreceptor cells, which varies widely from person to person, the model uses the relative proportion of rods and cones because these relative proportions are much more consistent across individuals. In doing so, the relative loss of rod and cones can be estimated by comparing ONL thickness to healthy controls. To demonstrate potential clinical utility of this model, we used it to estimate the degree of photoreceptor loss using a dataset of patients with molecularly confirmed STGD1. Leveraging retrospective OCT data acquired during routine clinical care, the model estimates survival of each photoreceptor cell type in STGD1 patients and reveals subtle loss that may not be readily apparent on qualitative analysis even by expert clinicians.

Rods and cones exhibit differential survival patterns across various forms of retinal degeneration, including age-related macular degeneration and rarer conditions such as inherited retinal diseases. Distinguishing between cone death and rod death can help the clinician narrow the range of considered diagnoses^20^, unravel mechanisms of progression^2^, and categorize patients by disease stage. Despite the importance of this problem, quantifying the degree of rod and cone loss *in vivo* remains a substantial challenge using available retinal imaging. Advances in adaptive-optics (AO) coupled with existing modalities, such as OCT, scanning laser ophthalmoscopy (SLO) or flood-illuminated ophthalmoscopy, have enabled quantification of photoreceptor cells. However, AO systems are expensive, not widely available, technically challenging to use, easily subject to artifacts such as patient movement, and typically restricted to visualizing a few degrees of the retina at a time. Moreover, commercially available AO systems can only reliably count cones but not rods^21-23^. Furthermore, due to optical limitations, these systems have difficulty resolving cones in the fovea, where cone density is highest and the most important for visual prognosis.

Compared to AO-imaging, OCT offers a wider field of view, faster acquisition, and greater familiarity among physicians and photographers due to its widespread use. The model proposed in this paper enables the estimation of rod and cone loss using readily available OCT data alone. An OCT-based computational model of photoreceptor loss has numerous advantages compared to AO, including its ability to analyze large, retrospectively collected OCT datasets acquired during routine clinical practice, the availability of longitudinal OCT data (in many cases over a decade or more) to compare changes over time, and clinician familiarity with this technology to facilitate interpretation of model data with structural information conveyed in the OCT B-scans (e.g., **Figure 4**).

Our model proposed here relies on several assumptions. We chose to build our model on the publicly available mean cell density data from Curcio and coworkers’ landmark 1990 study of retinal anatomy^1^. The accuracy of this dataset was recently confirmed using a larger cohort^7^ and imaged with a custom AO-SLO system^24^. This follow up study concluded that, while the average cell densities originally reported in Ref.^1^ are slightly higher than those measured by AO-SLO, especially within 300 μm of the fovea, the overall pattern of cell densities were highly concordant between the two studies^7^. It is possible that registration to other density datasets might yield different estimations of rod and cone loss. Our model could readily register to alternative datasets or incorporate variability in expected cellular composition as well as averages for a study.

As a first approximation of rod and cone loss for this model, we focused on two regions based on the ETDRS grid, the central subfield and the outer ring. Clinicians are familiar with using the ETDRS grid for evaluating macular pathologies since the grid was first introduced in 1991^15^. Cones predominate in the center subfield and rods predominate in the outer subfield, allowing these two regions to serve as biomarkers for cone and rod health, respectively. By excluding the inner ring covering the perifoveal region, we bypassed the region where the proportion of rods and cones is expected to vary the most between individuals. Future versions of our model could refine the areas beyond just the center subfield and outer ring. For example, instead of analyzing changes within ETDRS regions, which can be subject to floor effects^25^, our model could be extended to estimate expected rod and cone survival for all A-scans.

Our model also makes simple assumptions regarding the direct correlation between retinal thickness and proportion of rod and cone loss per region. However, the process of retinal degeneration is complex and nonlinear, involving multiple processes. Scarring and macular edema, for example, can increase retinal thickness, suggesting that obvious gliosis or edema are exclusion criteria for our model. Even apart from retinal disease, the gradual loss of cones in normal aging may result in paradoxical thickening of the retina. A recent study paired OCT with AO-SLO to compare the combined thickness of the HFL and ONL to photoreceptor density in a group of younger eyes (8 subjects, mean age 27.2 years) to a group of older eyes (8 subjects, mean age 56.2 years)^26^. That study indicated that, contrary to expectation, the combined HFL and ONL thickens with age, despite the loss of cones. (However, a different group, using flood illuminated AO, failed to observe a statistically significant loss of cone density in a comparison with similar ages and larger sample size^27^.) Neither of these studies, however, presented a null model of the expectation of ONL thickness lost under cell loss. For similar study designs, our model can be used to construct a null model of ONL thinning based exclusively on expected cell loss. Future versions of the model can also utilize larger datasets including from healthy control patients to make the estimates of photoreceptor loss more statistically robust.

In summary, we developed a computational model to estimate the proportion of surviving rods and cones using *in vivo* OCT data alone, without the need for advanced retinal imaging such as AO. Our model has many potential broad applications for research or clinical use. For example, future automated estimation of rod and cone loss could facilitate diagnosis of rod-or cone-specific diseases by OCT alone. Characterizing the rate and pattern of photoreceptor loss may help identify patient outliers within age classes of inherited retinal diseases such as STGD1 to provide new insights into pathophysiology or focus investigations of genetic disease modifiers.

## Data Availability

All de-identified data produced in the present study are available upon reasonable request to the authors.

## Funding

The University of Iowa Institute for Vision Research, The University of Iowa, Iowa City, Iowa and NIH/NEI grant (P30 EY025580).

## Data Availability

All de-identified data produced in the present study are available upon reasonable request to the corresponding author.

## Contributions

SSW and ICH conceived and designed the project. DBC, ICH, JLA, EMS contributed to acquisition of the data. APD, JLC, MM, CRF, DBC, SSW contributed to analysis and interpretation of the data. All authors contributed to drafting and critical revision of the manuscript, read and approved the final version, and agree to be accountable for all aspects of the work.

## Additional Information

The authors declare no competing interests.

## Acknowledgements

We thank Laura C. Whitmore, PhD, and Allan M. Andersen, MD, for providing feedback on the manuscript.

## Notes

### Competing Interest Statement

The authors have declared no competing interest.

### Funding Statement

This study was funded by The University of Iowa Institute for Vision Research, The University of Iowa, Iowa City, Iowa and by NIH/NEI grant (P30 EY025580).

### Author Declarations

The Institutional Review Board of the University of Iowa gave ethical approval for this work.

